# Belief in Conspiracy Theory about COVID-19 Predicts Mental Health and Well-being: A Study of Healthcare Staff in Ecuador

**DOI:** 10.1101/2020.05.26.20113258

**Authors:** Xi Chen, Stephen X. Zhang, Asghar Afshar Jahanshahi, Aldo Alvarez-Risco, Huiyang Dai, Jizhen Li, Verónica García Ibarra

**Affiliations:** University of Nottingham, Ningbo, China; University of Adelaide, Adelaide, Australia; Pontificia Universidad Católica del Perú (PUCP), Lima, Peru; Universidad de Lima, Lima, Peru; Tsinghua University, Beijing, China; Universidad Politécnica Estatal del Carchi, Turkan, Ecuador

**Keywords:** Coronavirus, 2019-nCoV, Mental health, Psychiatric identification, Latin America

## Abstract

**Background:** Social media are becoming hotbeds of conspiracy theories, which aim to give resolute explanations on the cause of COVID-19 pandemic. Yet, no research has investigated whether individuals’ belief in conspiracy theory about COVID-19 is associated with mental health and well-being issues. This association enables an assessable channel to identify and reach people with mental health and well-being issues during the pandemic.

**Objective:** We aim to provide the first evidence of belief in conspiracy theory regarding the COVID-19 virus as a predictor of the mental health and well-being of healthcare workers.

**Methods:** We conducted a survey of 252 healthcare workers in Ecuador from April 10 to May 2, 2020. We analyzed the data of distress and anxiety caseness with logistic regression and life and job satisfaction with linear regression.

**Results:** Among the sampled healthcare workers in Ecuador, 24.2% believed that the virus was developed intentionally in a lab; 32.54% experienced distress disorder, and 28.17% had anxiety disorder. Compared to healthcare workers who were not sure where the virus originated, those who believed the virus was developed intentionally in a lab were more likely to have distress disorder and anxiety disorder and had lower levels of job satisfaction and life satisfaction.

**Conclusions:** This paper identifies belief in a COVID-19 conspiracy theory as an important predictor of distress, anxiety, and job and life satisfaction of healthcare workers. It enables mental health services to better target and help mentally vulnerable healthcare workers during the ongoing COVID-19 pandemic.

## INTRODUCTION

During the COVID-19 pandemic, social media are populated with conspiracy theories—attempts to explain the ultimate causes of significant social events as secret plots by powerful and malicious groups [1, 2]. The most popular examples include: “the coronavirus was developed in a lab”; “people developed COVID-19 to destroy the governance of President Trump”; “Coronavirus is caused by 5G and is a form of radiation poisoning transmitted through radio waves” ; and “the coronavirus was Bill Gates’ attempt to take over the medical industry” [3-5]. Merely the last one has been mentioned 295,052 times across social media, broadcast, traditional media and online sites during one week in May 2020[6]. A national survey in UK found that approximately 50% of the population endorsed conspiracy theories to some degree[5].

Individuals’ belief in conspiracy theories has been linked to maladaptive personality traits[7], mental disorders and lower well-being[8]. However, no research has studied whether the conspiracy belief about COVID-19 is associated with mental health and well-being. This association is important because the specific COVID-19 conspiracy belief in social media is directly assessable and hence more useful to identify people with mental health and well-being issues during the pandemic. This paper explores a COVID-19 specific conspiracy belief that the coronavirus was developed intentionally in a lab as a predictor of individuals’ mental health and well-being during the pandemic. In particular, we focus on the mental health and well-being of healthcare workers – a prevalent and emergent issue during the COVID-19 pandemic [9]. The identification of COVID-19 conspiracy belief as a marker of mental health issues in healthcare workers uncovers a new channel for psychiatric screening and health communication [10], opening new avenues of research for medical informatics.

Previous research of COVID-19 has been primarily conducted in United States, China, and European countries, and there is a need for research on low-and-middle-income countries [11]. This study focuses on Ecuador where the COVID-19 crisis presents a particularly serious threat for healthcare workers, given the country’s scarce healthcare resources[12]. We surveyed healthcare workers in Ecuador from April 10 to May 2, 2020. During this period, there were 26,336 confirmed cases of COVID-19 and 1063 deaths, making the small country of Ecuador one of countries with the highest cases and death per capita in the world[13].

## METHODS

### Sample and Procedure

We conducted online survey with a convenient sample, which covered healthcare workers in both urban and rural areas. We approached 401 healthcare workers who worked in hospitals, clinics, first emergency responders, medical wards, nursing homes, dental clinics, and pharmacies in the 24 provinces of Ecuador. We received 252 completed surveys (response rate: 62.8%) from 54 healthcare facilities in 13 provinces (29 facilities in Carchi, 9 facilities in Quito, and 16 facilities from 11 other provinces). Therefore, our sample covered a wide range of provinces that varied in the severity of the COVID-19 crisis.

The ethical approval (20200322) was obtained from Tsinghua University. All participants provided their informed consent, participated voluntarily and could terminate the survey at any time. The survey was anonymous, and confidentiality of information was assured.

### Measures

We assessed the participants’ socio-demographic characteristics, including gender, age, educational level, marriage status, and their exercise hours per day during the past week. COVID-19 status was measured by asking “Are you infected by COVID-19?” (No; unsure; or Yes). We used a measure of conspiracy belief specific to COVID-19, asking participants “from what you’ve seen or heard, what do you think is most likely the origin of the coronavirus”: 1) It came about naturally; 2) It was developed intentionally in a lab (conspiracy belief); 3) It was most likely made accidentally in a lab; 4) I am not sure where the virus originated[14].

We used a brief measure of generalized anxiety disorder (GAD-7), which has been used broadly to measure anxiety[15]. GAD-7 consists of seven questions, with a cutoff of 10 or greater indicating cases of generalized anxiety disorder (α = .87). Psychological distress was measured with the six-item K6 screening scale (α = .90)[16], with a cutoff of 13 representing distress disorder caseness. We conducted logistic regression to analyze anxiety and distress caseness.

Following the example of previous research [17, 18], we used life satisfaction and job satisfaction to measure healthcare workers’ well-being. Life satisfaction was measured by the satisfaction with life scale with five items, including “In most ways my life is close to my ideal” (1 = strongly disagree, 7 = strongly agree; α = .81)[19]. Job satisfaction was measured with five items, including “I feel fairly satisfied with my present job” (1 = strongly disagree, 7 = strongly agree; α = .78)[20]. We conducted linear regression to analyze life satisfaction and job satisfaction.

## RESULTS

### Descriptive findings

Table 1 presents the descriptive findings of the sampled healthcare workers. Of the healthcare workers, 24.2% (61) believed that the virus was developed intentionally in a lab; 20.6% (52) believed that the virus came about naturally; 13.9% (35) believed that it was made accidentally in a lab; and the remaining 41.3% (104) were unsure where it originated.

**Table 1.**
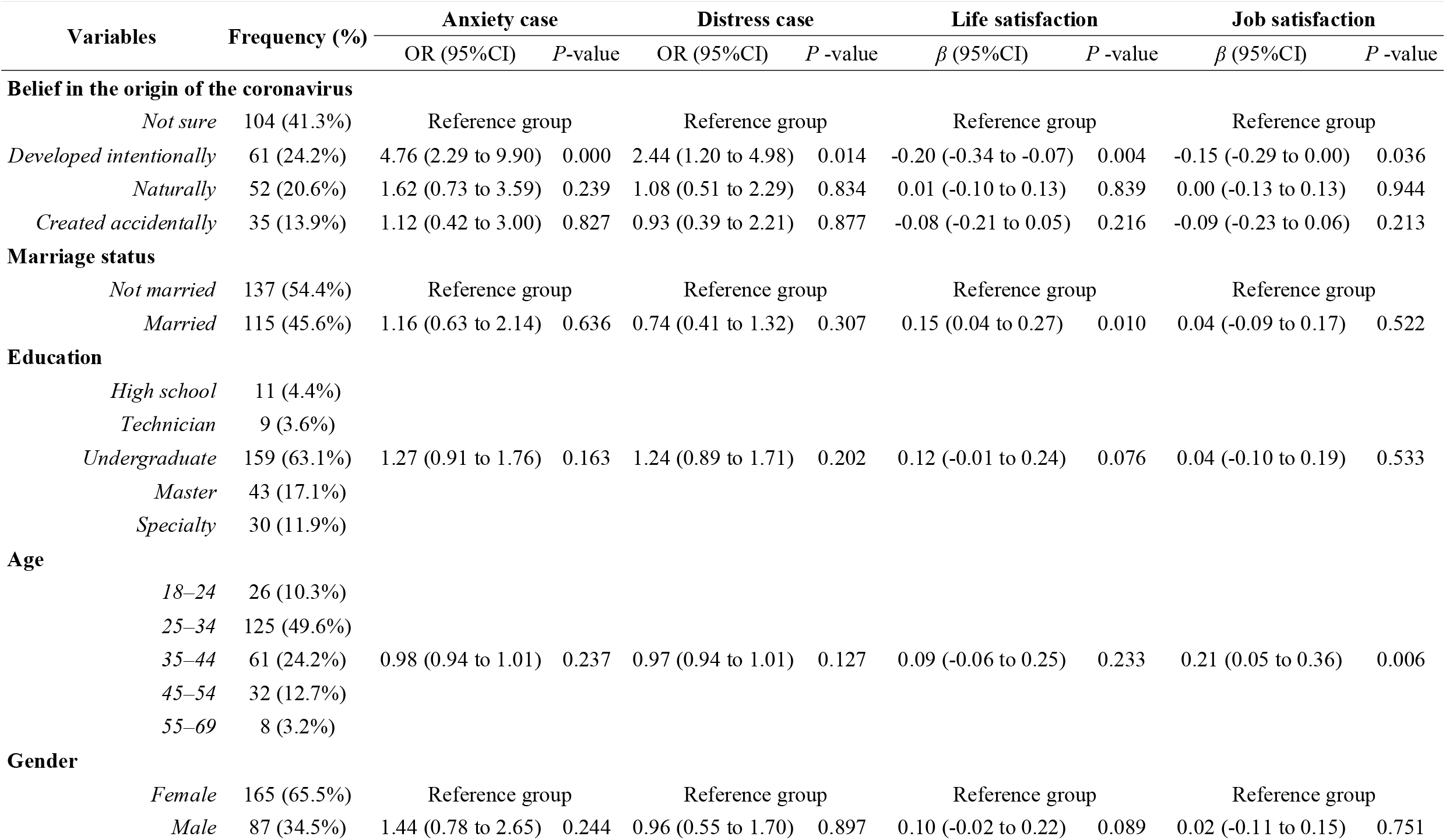
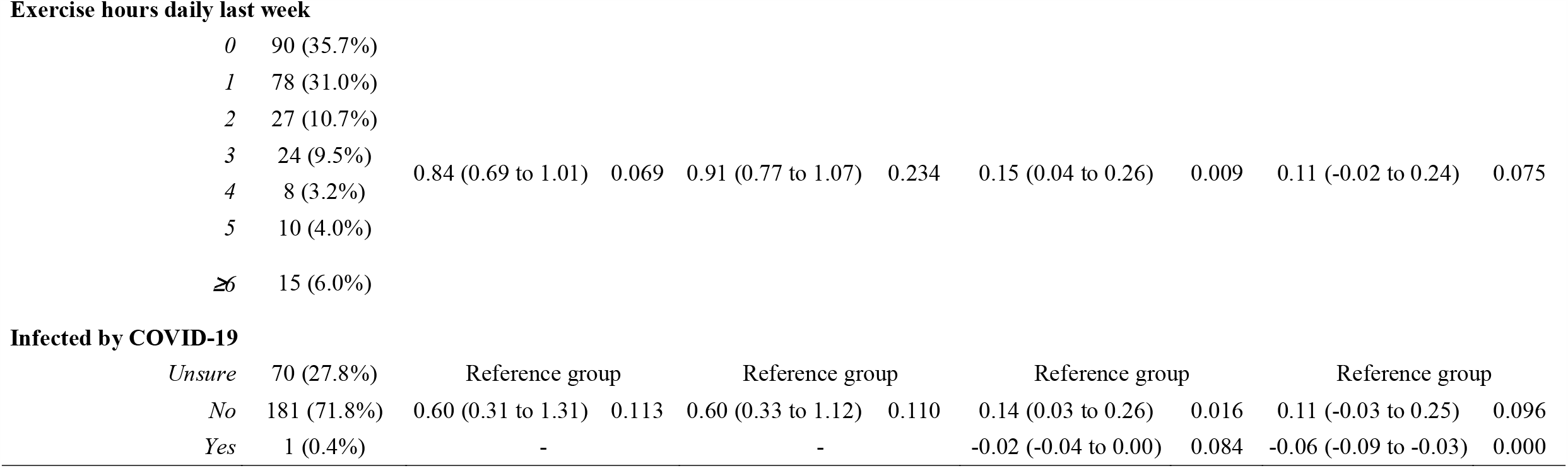
Descriptive findings and predictors of healthcare workers’ mental health and well-being by regression analyses (n= 252)

| Variables | Frequency (%) | Anxiety case |  | Distress case |  | Life satisfaction |  | Job satisfaction |  |
| --- | --- | --- | --- | --- | --- | --- | --- | --- | --- |
| | | OR (95%CI) | *P*-value | OR (95%CI) | *P* -value | $\beta$ (95%CI) | *P* -value | $\beta$ (95%CI) | *P* -value |
| Belief in the origin of the coronavirus |  |  |  |  |  |  |  |  |  |
| Not sure | 104 (41.3%) | Reference group |  | Reference group |  | Reference group |  | Reference group |  |
| Developed intentionally | 61 (24.2%) | 4.76 (2.29 to 9.90) | 0.000 | 2.44 (1.20 to 4.98) | 0.014 | -0.20 (-0.34 to -0.07) | 0.004 | -0.15 (-0.29 to 0.00) | 0.036 |
| Naturally | 52 (20.6%) | 1.62 (0.73 to 3.59) | 0.239 | 1.08 (0.51 to 2.29) | 0.834 | 0.01 (-0.10 to 0.13) | 0.839 | 0.00 (-0.13 to 0.13) | 0.944 |
| Created accidentally | 35 (13.9%) | 1.12 (0.42 to 3.00) | 0.827 | 0.93 (0.39 to 2.21) | 0.877 | -0.08 (-0.21 to 0.05) | 0.216 | -0.09 (-0.23 to 0.06) | 0.213 |
| Marriage status |  |  |  |  |  |  |  |  |  |
| Not married | 137 (54.4%) | Reference group |  | Reference group |  | Reference group |  | Reference group |  |
| Married | 115 (45.6%) | 1.16 (0.63 to 2.14) | 0.636 | 0.74 (0.41 to 1.32) | 0.307 | 0.15 (0.04 to 0.27) | 0.010 | 0.04 (-0.09 to 0.17) | 0.522 |
| Education |  |  |  |  |  |  |  |  |  |
| High school | 11 (4.4%) |  |  |  |  |  |  |  |  |
| Technician | 9 (3.6%) |  |  |  |  |  |  |  |  |
| Undergraduate | 159 (63.1%) | 1.27 (0.91 to 1.76) | 0.163 | 1.24 (0.89 to 1.71) | 0.202 | 0.12 (-0.01 to 0.24) | 0.076 | 0.04 (-0.10 to 0.19) | 0.533 |
| Master | 43 (17.1%) |  |  |  |  |  |  |  |  |
| Specialty | 30 (11.9%) |  |  |  |  |  |  |  |  |
| Age |  |  |  |  |  |  |  |  |  |
| 18–24 | 26 (10.3%) |  |  |  |  |  |  |  |  |
| 25–34 | 125 (49.6%) |  |  |  |  |  |  |  |  |
| 35–44 | 61 (24.2%) | 0.98 (0.94 to 1.01) | 0.237 | 0.97 (0.94 to 1.01) | 0.127 | 0.09 (-0.06 to 0.25) | 0.233 | 0.21 (0.05 to 0.36) | 0.006 |
| 45–54 | 32 (12.7%) |  |  |  |  |  |  |  |  |
| 55–69 | 8 (3.2%) |  |  |  |  |  |  |  |  |
| Gender |  |  |  |  |  |  |  |  |  |
| Female | 165 (65.5%) | Reference group |  | Reference group |  | Reference group |  | Reference group |  |
| Male | 87 (34.5%) | 1.44 (0.78 to 2.65) | 0.244 | 0.96 (0.55 to 1.70) | 0.897 | 0.10 (-0.02 to 0.22) | 0.089 | 0.02 (-0.11 to 0.15) | 0.751 |
| **Exercise hours daily last week** |  |  |  |  |  |  |  |  |  |
| *0* | 90 (35.7%) |  |  |  |  |  |  |  |  |
| *1* | 78 (31.0%) |  |  |  |  |  |  |  |  |
| *2* | 27 (10.7%) |  |  |  |  |  |  |  |  |
| *3* | 24 (9.5%) | 0.84 (0.69 to 1.01) | 0.069 | 0.91 (0.77 to 1.07) | 0.234 | 0.15 (0.04 to 0.26) | 0.009 | 0.11 (-0.02 to 0.24) | 0.075 |
| *4* | 8 (3.2%) |  |  |  |  |  |  |  |  |
| *5* | 10 (4.0%) |  |  |  |  |  |  |  |  |
| *≥6* | 15 (6.0%) |  |  |  |  |  |  |  |  |
| **Infected by COVID-19** |  |  |  |  |  |  |  |  |  |
| *Unsure* | 70 (27.8%) | Reference group |  | Reference group |  | Reference group |  | Reference group |  |
| *No* | 181 (71.8%) | 0.60 (0.31 to 1.31) | 0.113 | 0.60 (0.33 to 1.12) | 0.110 | 0.14 (0.03 to 0.26) | 0.016 | 0.11 (-0.03 to 0.25) | 0.096 |
| *Yes* | 1 (0.4%) | - |  | - |  | -0.02 (-0.04 to 0.00) | 0.084 | -0.06 (-0.09 to -0.03) | 0.000 |

### Predictors of healthcare workers’ mental health

As presented in Table 1 and further illustrated in Figure 1 in the online supplement, healthcare workers who believed that the virus was developed intentionally in a lab were more likely to experience distress disorder than those who were unsure of the origin of the virus. Wald test showed that they were also more likely to experience distress disorder than those who believed the virus was created accidentally (χ^2^ (1) = 4.24, *P* = 0.039).

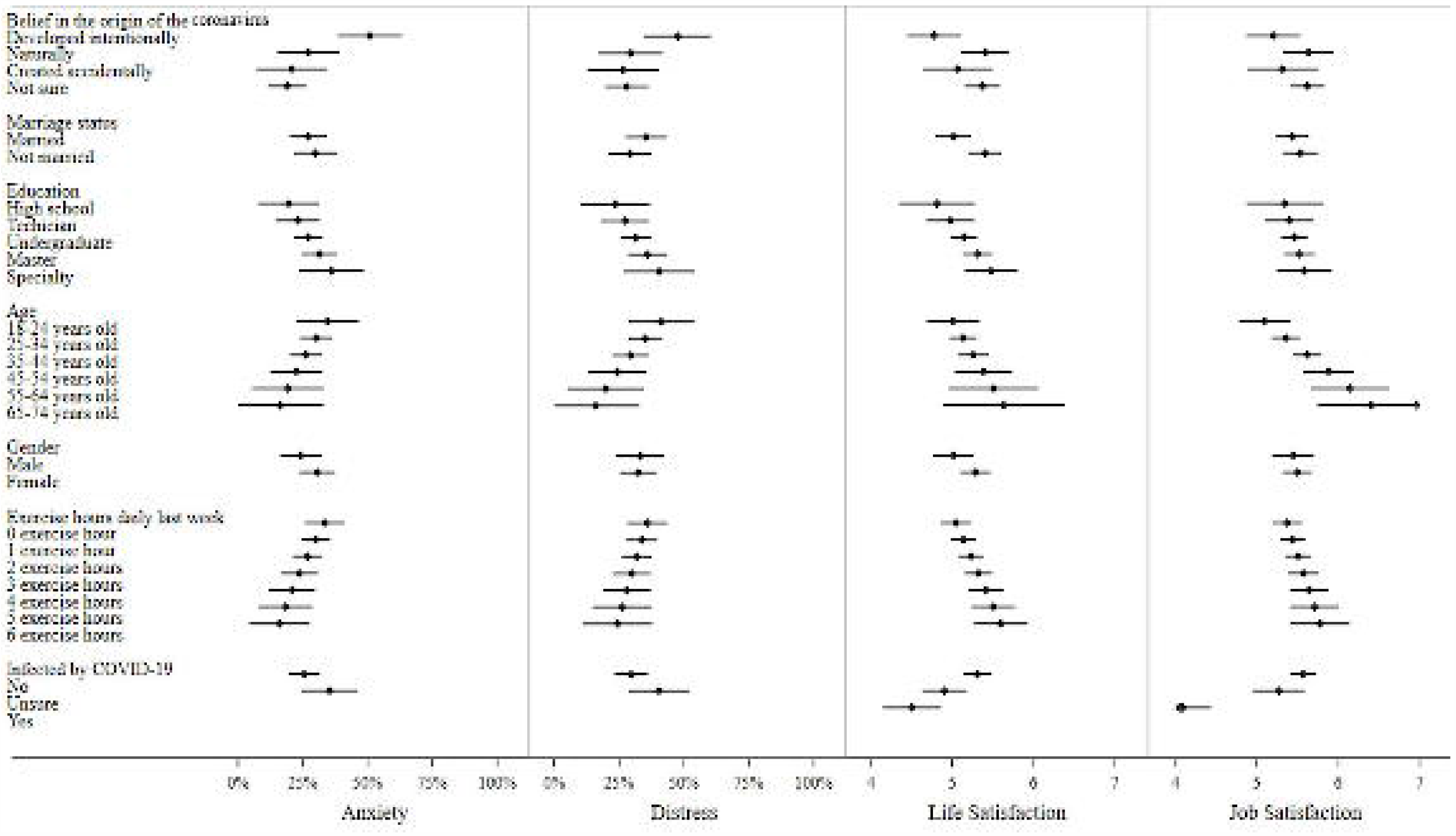

Healthcare workers who believed the virus was developed intentionally in a lab were more likely to have anxiety disorder than those who were unsure how the virus originated. Wald test showed that they were also more likely to have anxiety disorder than those who believed the virus came about naturally (χ^2^ (1) = 6.42, *P* = 0.011) and those who believed the virus was made accidentally (χ^2^ (1) = 8.11, *P* = 0.004).

### Predictors of healthcare workers’ well-being

Healthcare workers who were married or exercised more hours in the past week had higher life satisfaction. Those who were not affected by COVID-19 were more satisfied with life than those who were unsure. Healthcare workers who viewed the virus as developed intentionally in a lab had lower life satisfaction than those who were unsure how the virus originated. Wald test showed that their life satisfaction was also lower than those who believed the virus came about naturally (χ^2^ (1) = 7.80, *P* = 0.006).

Older healthcare workers had higher job satisfaction. Healthcare workers who believed the virus was developed intentionally in a lab had lower job satisfaction than those who were unsure how the virus originated.

## DISCUSSION

First, this study revealed that even healthcare professionals could believe in conspiracy theories (24.2% in this sample). The prevalent belief in conspiracy theory is related to the high anxiety and distress of healthcare workers in Ecuador. Almost one third (32.5%) of the healthcare workers surpassed the cutoff of distress disorder, and 28.2% had anxiety disorder. The proportion of distressed healthcare workers in Ecuador was significantly higher than healthcare workers in Iran surveyed on February 28–30, 2020 (20.1%, N = 304)[21]. The prevalence of anxiety disorder was similar to a sample of 5062 healthcare workers (24.06% by the cutoff of 8) in Wuhan of China during February 8–10, 2020 [22] and higher than a sample of 4872 individuals (22.6% by the cutoff at 10) in China surveyed during January 31 to February 2, 2020[23].

This study found that conspiracy belief in the origin of COVID-19 was associated with lower mental health, life satisfaction, and job satisfaction of healthcare workers. From a health informatics perspective, the belief in a COVID-19 related conspiracy theory provides a marker to identify mentally vulnerable people, who may browse, search, follow, like, discuss, and disseminate COVID-19 related conspiracy theories via social media and other channels. Such information can serve as a risk factor to identify individuals more susceptible to mental disorders in psychiatric screening via social media[24], at a time when the psychological screening, diagnosis, and intervention are rapidly moving online [25].

In addition, this study also provides important implications for the dissemination of scientific and health information. Previous research has recognized the important role of online scientific communication in combating conspiracy theories [1, 26]. This study suggests that such communication should acknowledge recipients’ psychological states, such as anxiety and distress, while introducing scientific hypotheses about the origin of the virus [27]. Given that believers of conspiracy theories tend to cluster[4], the followers of COVID-19 related conspiracy theories also provide targeted groups for scientific communications and mental health information dissemination [10].

Finally, the conspiracy belief that the virus was developed intentionally in a lab was associated with reduced job satisfaction of healthcare workers. Given that the mental health of healthcare workers is important to sustain their employment and job performance[28], this study highlights the important role of conspiracy theories in detecting healthcare staff’s mental health, which has profound implications for their overall performance. This is especially important in settings where healthcare resources are already constrained, such as the COVID-19 pandemic.

### Limitations and future research

There are several limitations of this study. First, the cross-sectional design limits our ability to make causal arguments about the relationship between conspiracy belief and mental health. Future research should adopt experimental designs to establish the causal relationship between conspiracy belief and mental health. Second, we only focused on healthcare workers, whose role is especially important during the ongoing COVID-19 pandemic in Ecuador. It is worth investigating if the effects of conspiracy belief generalize to the general population. Finally, Ecuador is a country that is suffering a serious toll from the pandemic. It remains to be seen to what extent the findings are generalizable to other countries, which face different degrees of threat from the pandemic. For instance, it may be interesting to investigate if conspiracy theory about COVID-19 predicts mental health in countries where the social and political systems are severely threatened by the pandemic, because system threat is an important cause of adopting conspiracy theory[29].

## Conclusion

In conclusion, this study provides the first empirical evidence that COVID-19 related conspiracy belief was associated with mental health and well-being of healthcare workers. Hence, belief in COVID-19 related conspiracy theories expressed on social media and interest groups may help identify mentally vulnerable people to enable more targeted identification and communication from a health informatics perspective.

## Data Availability

Available upon request.

## Acknowledgement

We acknowledge the support of Tsinghua University-INDITEX Sustainable Development Fund (Project No. TISD201904).

